# Telomere maintaining germline and somatic variants in thyroid cancer and melanoma

**DOI:** 10.64898/2026.05.22.26353814

**Authors:** Sandya Liyanarachchi, Pamela L. Brock, Wei Li, Taina T Nieminen, Nikita Pozdeyev, Bryan R Haugen, Hilary C. McCrary, Bodour Salhia, Kirk Jensen, Abdul Rafeh Naqash, Varinder Kaur, Janice Farlow, Matthew D. Ringel

**Affiliations:** Department of Molecular Medicine and Therapeutics, The Ohio State University Comprehensive Cancer Center, Columbus, Ohio, USA; Division of Human Genetics, The Ohio State University College of Medicine, Comprehensive Cancer Center, Columbus, Ohio, USA; Department of Medical and Clinical Genetics, University of Helsinki, Helsinki, Finland; Department of Biomedical Informatics, University of Colorado Anschutz, Aurora, CO, USA. Division of Endocrinology, Diabetes and Metabolism, Colorado Center for Personalized Medicine, University of Colorado Cancer Center, University of Colorado Anschutz, Aurora, CO, USA; Division of Endocrinology, Diabetes and Metabolism, University of Colorado Cancer Center, University of Colorado Anschutz Medical Campus, Aurora, CO, USA; Department of Otolaryngology-Head and Neck Surgery University of Utah School of Medicine and Huntsman Cancer Institute, Salt Lake City, UT, USA; Department of Translational Genomics, Keck School of Medicine, University of Southern California and Norris Comprehensive Cancer Center, Los Angeles, CA, USA; Uniformed Services University of the Health Sciences, Bethesda, MD; Stephenson Cancer Center, University of Oklahoma Health Sciences Center, Oklahoma City, Oklahoma, USA; Department of Internal Medicine, Division of Hematology & Oncology, University of Virginia School of Medicine and University of Virginia Cancer Center, Charlottesville, VA, USA; Indiana University School of Medicine, Indianapolis, IN, USA; Departments of Molecular Medicine and Therapeutics and Internal Medicine, The Ohio State University College of Medicine and Comprehensive Cancer Center, Columbus, Ohio, USA

## Abstract

**Importance:** Non-medullary thyroid cancer (NMTC) and melanoma are associated with inherited long telomeres due to germline pathogenic/likely pathogenic variants (PV/LPV) in *POT1*, *TINF2*, and *ACD* resulting in long-telomere syndrome (LTS) and they commonly have somatic *TERT* promoter mutations. The genetic relationship between these variants and their clinical associations are defined incompletely and may inform clinical practice.

**Objective:** To test the hypothesis that germline LTS-associated PV/LPV are exclusive from functional somatic *TERT* variants and assess clinical/genetic associations.

**Design:** Retrospective observational cohort study with/without germline LTS variants, that have somatic sequencing and pathology data.

**Setting:** Participants were enrolled through 18 cancer centers participating in the Oncology Research Information Exchange Network (ORIEN).

**Participants:** 995 adults with NMTC and 993 with melanoma between 2013 and 2025. All adult patients at an ORIEN center were offered enrollment

**Exposures:** All patients with NMTC or melanoma are included. There are no required exposures.

**Main Outcomes and Measures:** The presence/absence of a germline or somatic long-telomere variant; secondary outcomes are associations with tumor stage, telomerase expression, and oncogenes.

**Results:** Germline and somatic variants in *POT1*/*TINF2/ACD*, somatic *TERT* promoter variants, *TERT* fusions, oncogenes, and telomerase mRNA expression were evaluated in 995 NMTC and 993 melanoma patients. In NMTC, 13 (1.5%) had a germline LTS variant while 0/12 with tumor sequencing had somatic *TERT* promoter variants/fusions. In melanoma, 7 (0.7%) had a LTS variant; 0/2 with tumor sequencing had a *TERT* promoter variant/ fusion. Meta-analysis including NMTC and melanoma in the current study, a recent thyroid cancer study, and thyroid TCGA, germline LTS-associated PV/LPV and somatic *TERT* variants/fusions were mutually exclusive (p=0.036). High telomerase mRNA levels were associated with *TERT* promoter variants/fusions (p<4e-11) and larger NMTC/distant metastases (p=0.016), but not germline LTS variants. NMTCs with somatic *TERT* promoter variants/fusions had higher tumor mutation burden (p<0.02) versus tumors from patients with a germline LTS variant. *TERT* promoter mutant variant allele frequency was lower in smaller and non-metastatic vs larger/metastatic NMTC.

**Conclusion and Relevance:** Germline LTS-associated variants appear to be exclusive from somatic *TERT* promoter variants/fusions but are not associated with aggressive NMTC, suggesting common roles in tumorigenesis but different biological impacts.

**Key points:** *Question:* Are germline and somatic variants associated with telomere maintenance mutually exclusive and are their biological associations distinct or overlapping?

*Findings:* In this cohort study, it was determined that germline variants that cause long telomere syndrome are mutually exclusive from somatic *TERT* promoter mutations in non-medullary thyroid cancer (NMTC) and melanoma but that the somatic *TERT* promoter mutations are uniquely associated with more aggressive disease in NMTC.

*Meaning:* Despite the mutual exclusivity of germline and somatic telomere maintenance variants in patients with thyroid cancer, the clinical associations are not overlapping, consistent with functional differences supporting the need for individualized clinical counseling.

## INTRODUCTION

Individuals with germline pathogenic and likely pathogenic variants (PV/LPVs) leading to long telomeres have an increased risk for clonal hematopoiesis, certain malignancies, and possibly benign tumors^1–3^. This emerging predisposition condition has been termed “long telomere syndrome (LTS)”^4^. Variants in three genes (*ACD, POT1*, and *TINF2*), which encode telomere-binding proteins, have been identified as causative for LTS^1,5^. Individuals with PV/LPVs in these genes typically maintain ultra-long telomere length maintained over time. This presumably leads to delayed senescence and altered DNA damage responses, with the acquisition of mutations increasing the risk of malignancy. In addition, affected families have demonstrated progressive telomere elongation in successive generations, suggesting genetic anticipation^2^.

Increased cancer incidence has been associated with either short or long telomeres^6–8^. The malignancies most associated with LTS include melanoma, leukemia, papillary thyroid cancer (PTC), sarcoma, and others. In one study, among 17 individuals with germline *POT1* variants and long telomere length, PTC was the second most common solid tumor^2^. This led to our prior investigation of a larger cohort of patients with PTC. In the overall group of 470 individuals with PTC, LPV/PVs in *ACD, POT1*, and *TINF2* were found in 4.5% and 1.5% of familial and unselected cases, respectively. Affected individuals had ultra-long telomere length, and the majority (83%) had multiple cancers, including PTC, melanoma, lymphoma, and sarcoma. The strongest co-occurrence relationship was between PTC and melanoma; 22% of individuals with both diagnoses had a germline LTS-associated variant ^1^. In addition, at least three families with autosomal dominant inherited PTC (in association with other cancers) linked to germline PVs in *TINF2* and *POT1* have been reported^5,9,10^. These data suggest a potentially important relationship between inherited long telomeres and PTC and melanoma.

Somatic mutations in the *TERT* promoter are common in PTC and melanoma. In non-medullary thyroid cancer (NMTC), the combination of *TERT* promoter mutations with *BRAF, RAS* activating mutations, or *RET* fusions is associated with aggressive disease and poor prognosis^11–15^. It has been shown that the two most common *TERT* promoter mutations at positions −128 and −250 result in new ETS2 binding sites which, in the context of MAPK activation and other pathways, result in *TERT* overexpression^16–18^. It has been proposed that *TERT* promoter mutations are secondary events that lead to aggressive disease in NMTC^19,20^. However, mutations in the *TERT* promoter can occur in isolation in benign thyroid nodules and occur early in melanoma, suggesting a potential role in facilitating NMTC development ^21,22^.

To begin to understand the relationship between germline telomere lengthening and somatic variants, we reported somatic data from 10 PTC samples from patients with germline variants in *ACD, POT1*, and *TINF2*. In this small cohort, all PTCs were negative for *TERT* promoter mutations and were positive for *BRAF V600E*^1^. Given these findings, we hypothesized that germline LTS PV/LPVs and somatic *TERT* promoter mutations or *TERT* fusions are mutually exclusive in a larger unselected cohort of patients with NMTC or melanoma. In addition, we wished to determine the clinical associations of the two long telomere mechanisms.

## METHODS

Germline and tumor whole exome and RNA sequencing were performed on blood and tumor specimens from 995 adults with NMTC and 993 adults with melanoma who consented to the Total Cancer Care protocol at one of 18 cancer centers nationwide participating in the Oncology Research Information Exchange Network (ORIEN) between 2013 and 2025. The age at diagnosis ranged from 9 to 88 years for NMTC and 16 to 90 years for melanoma. All protocols were approved by the respective Institutional Review Boards, including OSU protocol 2013H0197.

### Whole exome sequencing data analysis

Whole exome sequencing (WES) samples were processed and analyzed as described in Brock et al^23^ with mean targeted coverage >50X and over 80% of bases having 20X or more coverage to pass quality thresholds. Somatic variants were filtered to have a minimum read coverage of 20 and a minimum variant allele frequency of 5%.

Variants were annotated using Variant Effect Predictor (VEP) v115.1 with genome build GRCh38 to estimate pathogenicity^24^. Germline variants in genes previously associated with LTS (*ACD*, *POT1*, and *TINF2*) were considered PV/LPV if they had at least one PV or LPV classification in ClinVar, if they were truncating or frameshift (without evidence of nonsense mediated decay escape), if they were missense with 2 out of 3 (CADD > 25, REVEL > 0.6, MetaRNN > 0.8), or if they were splice region variants +/− 1, 2 or 3 from intron/exon border with a max SpliceAI score > 0.6.

Three hybridization capture kits were utilized in ORIEN including NimbleGen (NIM), Integrated DNA Technologies (IDT), and Twist Bioscience (TWIST). There was very low coverage of the *TERT* promoter by the NIM capture kit; therefore, *TERT* promoter mutation status is unknown for these samples (**eFigure 1**). For samples with *TERT* promoter coverage, variant allele frequency (VAF) of *TERT* promoter mutations was estimated as the ratio between read counts of altered and total alleles.

### RNA sequencing data analysis

RNA sequencing samples were processed with Illumina TruSeq using the STAR (v.2.7.3a) aligner with human genome reference (GRCh38/hg38) and Gencode genome annotation v32. Quality control (QC) measures: duplicate rate < 90%, passing filter (PF) reads mapping to the targets over 50 million (> 50M), exonic rate >0.6 and percentage of RNA fragments that are longer than 200 nucleotides > 20% were used to filter out low-quality RNA sequencing samples. Gene fusions (specifically those involving *TERT*) were identified by analyzing RNA-Sequencing data applying STAR-Fusion (v1.8.0) and Arriba (v1.1.0) methods. Variant stabilized (VST) normalized gene expression estimates were obtained by applying the DESeq2 method^25^ correcting for sequencing depth and gene-wise variability using RNA sequencing read count data. Principal component analysis was performed with VST-normalized gene expression data. Batch correction to adjust for the differences in preservation methods was applied. For the 6 patients with multiple RNASeq data available, average gene expression estimates were used for analysis.

#### TaqMan assay analysis

*TERT* expression in 36 primary tumor and 21 metastatic tumor samples not included in ORIEN were measured TaqMan RTqPCR (Thermo Fisher *TERT* TaqMan assay Hs00972650_m1). *TERT* C228T/C250T mutations and the presence or absence of *BRAF* V600E in these samples were assessed by Sanger sequencing.

#### TCGA THCA data analysis

A similar analysis as noted above was performed using TCGA (https://www.cancer.gov/ccg/research/genome-sequencing/tcga) thyroid cancer (THCA) data^26^. STAR counts were downloaded using TCGAbiolinks package in R^27^ and VST normalized gene expression estimates were obtained by applying DeSeq2 package. Protected TCGA-THCA germline WES data was analyzed to identify germline LPV/PV in *ACD*, *POT1* and *TINF2*^25^. Protected TCGA-THCA somatic tumor WGS data was analyzed to identify *TERT* promoter mutation status and the supplemental data was used to extract the *TERT* status for the missing WGS samples^28^.

### Statistical analysis

Analyses were conducted with R v4.5.2 and RStudio 2026.01.0+392, as described in the **eMethods**.

## RESULTS

There were 995 NMTC and 993 melanoma patients with at least one type of sequencing data (germline WES, tumor WES, or RNASeq) available after applying quality filters. Demographic and clinical information for both cohorts are displayed in **Table 1**.

**Table 1.** ORIEN NMTC and melanoma cohort features.

|  | **ORIEN NMTC (n=995)** | **ORIEN Melanoma (n=993)** |
| --- | --- | --- |
| **Age, mean ± std** | 47.84 ± 15.96 | 59.16 ± 14.33 |
| **Sex, n (%)** |  |  |
| Male | 359 (36.08%) | 615 (61.93%) |
| Female | 636 (63.92%) | 378 (38.07%) |
| **Race, n (%)** |  |  |
| AIAN | 4 (0.40%) | 1 (0.10%) |
| Asian | 40 (4.02%) | 2 (0.2%) |
| Black | 26 (2.61%) | 3 (0.3%) |
| NHPI | 4 (0.40%) | 0 (0.00%) |
| White | 906 (91.06%) | 969 (97.58%) |
| Other | 12 (1.21%) | 6 (0.60%) |
| Unknown | 3 (0.30%) | 12 (1.21%) |
| **Histology, n (%)** |  |  |
| PTC | 734 (73.77%) | N/A |
| FVPTC | 164 (16.48%) | N/A |
| FTC | 51 (5.13%) | N/A |
| OTC | 36 (3.62%) | N/A |
| PDTC | 4 (0.40%) | N/A |
| NIFTP | 6 (0.6%) | N/A |
| **T, n (%)** |  |  |
| T0 | 2 (0.20%) | 40 (4.03%) |
| T1 | 355 (35.68%) | 102 (10.27%) |
| T2 | 230 (23.12%) | 91 (9.16%) |
| T3 | 303 (30.45%) | 138 (13.90%) |
| T4 | 60 (6.03%) | 193 (19.44%) |
| Tis | 2 (0.20%) | 68 (6.85%) |
| Unknown | 43 (4.32%) | 363 (36.35%) |
| **N, n (%)** |  |  |
| N0 | 429 (43.12%) | 342 (34.44%) |
| N1 | 462 (46.43%) | 111 (11.18%) |
| N2 | N/A | 79 (7.96%) |
| N3 | N/A | 74 (7.45%) |
| Unknown | 104 (10.45%) | 389 (38.97%) |
| **M, n (%)** |  |  |
| M0 | 834 (83.82%) | 567 (57.10%) |
| M1 | 36 (3.62%) | 88 (8.86%) |
| Unknown | 125 (12.56%) | 340 (34.04%) |

AIAN=American Indian or Alaska Native, NHPI=Native Hawaiian or Pacific Islander, N/A=Not Applicable

### ORIEN NMTC and Melanoma Drivers and Populations

Among the 995 NMTC patients, 971 patients had WES of a tumor sample (891 primary tumor, 166 metastasis, and 62 from both) and 649 patients had RNASeq (574 primary tumor, 129 metastasis, and 54 from both). Among the 993 melanoma patients, 593 patients had WES of a tumor sample (199 primary tumor, 415 metastasis, and 21 from both) and 443 patients had RNASeq (145 primary tumor, 308 metastasis, and 10 from both). **eFigure 2** and **eFigure 3** depict the distributions of the NMTC and melanoma cohort with respect to tumor *BRAF/RAS* variant and *TERT* promoter status, and availability of *TERT* expression data, respectively.

### Germline Variants

In the NMTC cohort, 894 patients had germline exome data available. Of these, 13 (1.5%) had PV/LPV variants in one of the 3 genes associated with LTS, including 3 in *ACD*, 6 in *POT1*, and 4 in *TINF2*. For the 101 patients with no germline exome data, somatic exome data and/or RNASeq data were assessed and no potential germline variants were identified. Among the 13 NMTC patients with germline *ACD*, *POT1*, or *TINF2* PV/LPV, 12 had PTC and one had poorly differentiated thyroid cancer (PDTC). Twelve (12/13 or 92.3%) of the germline positive cases had a somatic activating *BRAF* (10/13) or *RAS* variant (2/13), with the remaining case having an activating *BRAF* variant at a low VAF (1.5%). None of the 12 cases with *TERT* promoter coverage had a *TERT* promoter variant.

When compared to frequency of somatic *TERT* promoter mutations in patients without LTS germline variants (102/640 or 15.9%), patients with germline variants were more likely to be negative for *TERT* promoter mutations (OR= 4.97, p= 0.117, **Table 2a**), based on OR; p values were likely influenced by small number of patients with germline variants.

**Table 2.**
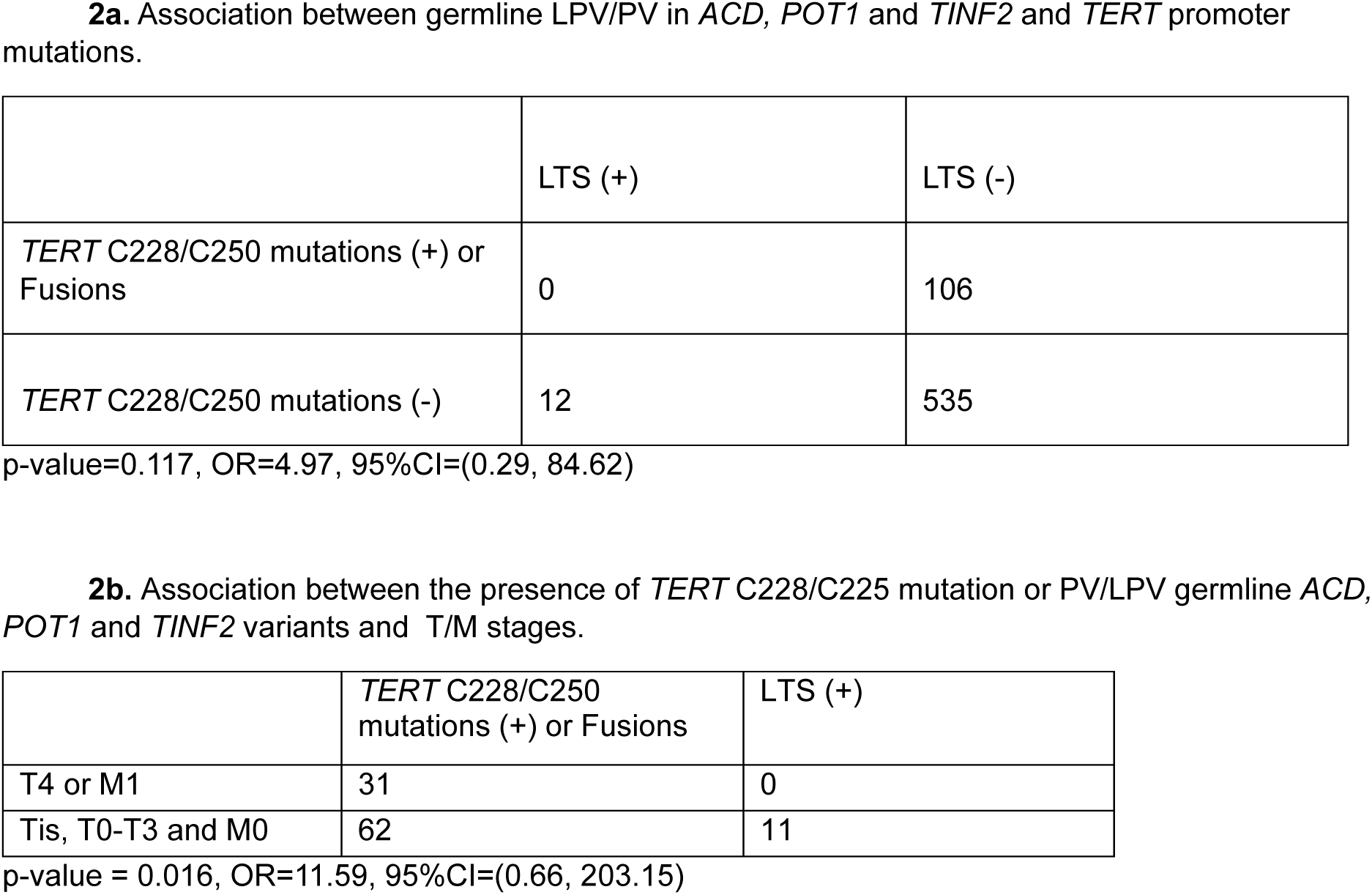
Associations between germline LPV/PV in *ACD, POT1* and *TINF2*, and *TERT* promoter mutations, and T/M stages in the ORIEN NMTC cohort.

Five of these 13 cases overlapped with the 10 PTC patients reported previously in Deboy et al^1^. Three patients in the thyroid TCGA cohort also had LTS germline variants. **eTable 1** provides histology and variant information for patients with germline *ACD*, *POT1*, or *TINF2* PV/LPV among the ORIEN, DeBoy, et al, and TCGA-THCA cohorts. In combination there are 20 patients with germline LTS variants. None have somatic *TERT* promoter mutations, significantly lower than LTS negative patients from those cohorts with both germline and somatic data, (0/20 (0%) vs 159/971 (16.4%); p=0.0496).

Most LTS germline carriers had tumors with *BRAF* V600E or *RAS/RAS*-like mutations; however, the enrichment did not reach statistical significance versus the LTS negative population [92.3% vs 65.9%; p=0.072 and OR=6.21 (95% CI=0.91-266.1)] (**Figure 1a**). The proportion of patients with T4 or M1-stages was higher for those with somatic *TERT* promoter mutations or fusions versus those without (33.33% vs 4.82%, p =3.00e-13; OR=9.80; 95%CI=5.13, 19.00) (**eTable 2**). We observed that none of the primary NMTC patients with LTC variants were diagnosed with T4 or M1 stages while 33.33% of patients with *TERT* promoter mutations/fusions were diagnosed with T4 or M1 stages (p=0.016, OR=11.59; 95%CI=0.66, 203.15) (**Table 2b**).

**Figure 1.**
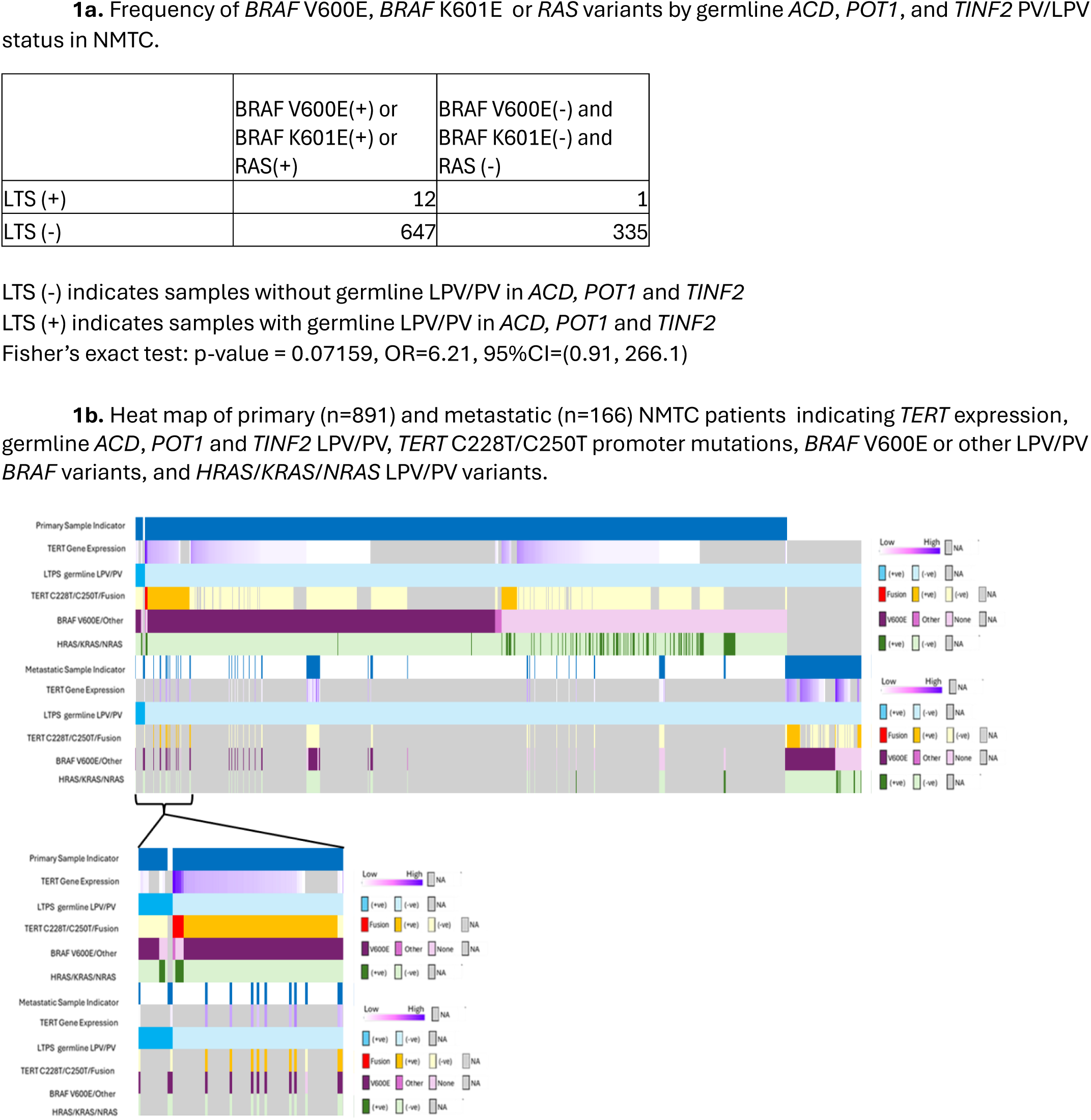
Oncoplot and frequency distribution. 1a: Frequency distribution of *BRAF* V600E, *BRAF* K601E or *RAS* variants by germline *ACD*, *POT1*, and *TINF2* PV/LPV status in NMTC, p=0.07159, OR=6.21, 95%CI=(0.91, 266.1). 1b: Oncoplot of primary (n=891) and metastatic (n=166) NMTC patients indicating *TERT* expression, germline *ACD*, *POT1* and *TINF2* LPV/PV, *TERT* C228T/C250T promoter mutations, *BRAF* V600E or other LPV/PV *BRAF* variants, and *HRAS/KRAS/NRAS* LPV/PV variants.

Of the 993 patients in the melanoma cohort with WES data, 7 (0.7%) patients had LTS LPV/PV variants including one in *ACD* and 6 in *POT1* (**eTable 1**). Among the 7 patients with germline *ACD* or *POT1* LPV/PV, two had tumor sequencing available and both were negative for *TERT* promoter mutations or *BRAF* V600E, and one had a somatic *NRAS* variant.

In combination, *TERT* promoter variants or fusions were absent from all 14 ORIEN patients with NMTC or melanoma and germline LTS variants. In the ORIEN germline negative cohorts, *TERT* promoter variants or fusions were more common (403/1111 or 36.27%, p=0.0019). A meta-analysis across the thyroid and melanoma cohorts confirmed a significant mutually exclusive relationship between germline LTS variants and somatic *TERT* promoter variants/fusions (p=0.036).

### *TERT* Fusions

*TERT* expression analysis of the NMTCs revealed four cases with markedly elevated *TERT* expression; the three evaluable cases were negative for *TERT* promoter mutations. Further investigation identified *TERT* fusions in all four cases (**Figure 2a**) including a *PDE8B:TERT* fusion in three samples and a novel *TERT* fusion (*NCOA3:TERT*) in one case. All samples had either a *RAS* or *RAS*-like *BRAF* variant (K601E). Similar analysis of the melanoma cohort identified four cases with *TERT* fusions with *PPP1R12A*, *LAPTM4B*, *SLC6A19*, and *EXOC3-AS1* (**Figure 2a**). All four were negative for *TERT* promoter mutations, one case was positive for *BRAF* V600E and one for an *NRAS* variant.

**Figure 2.**
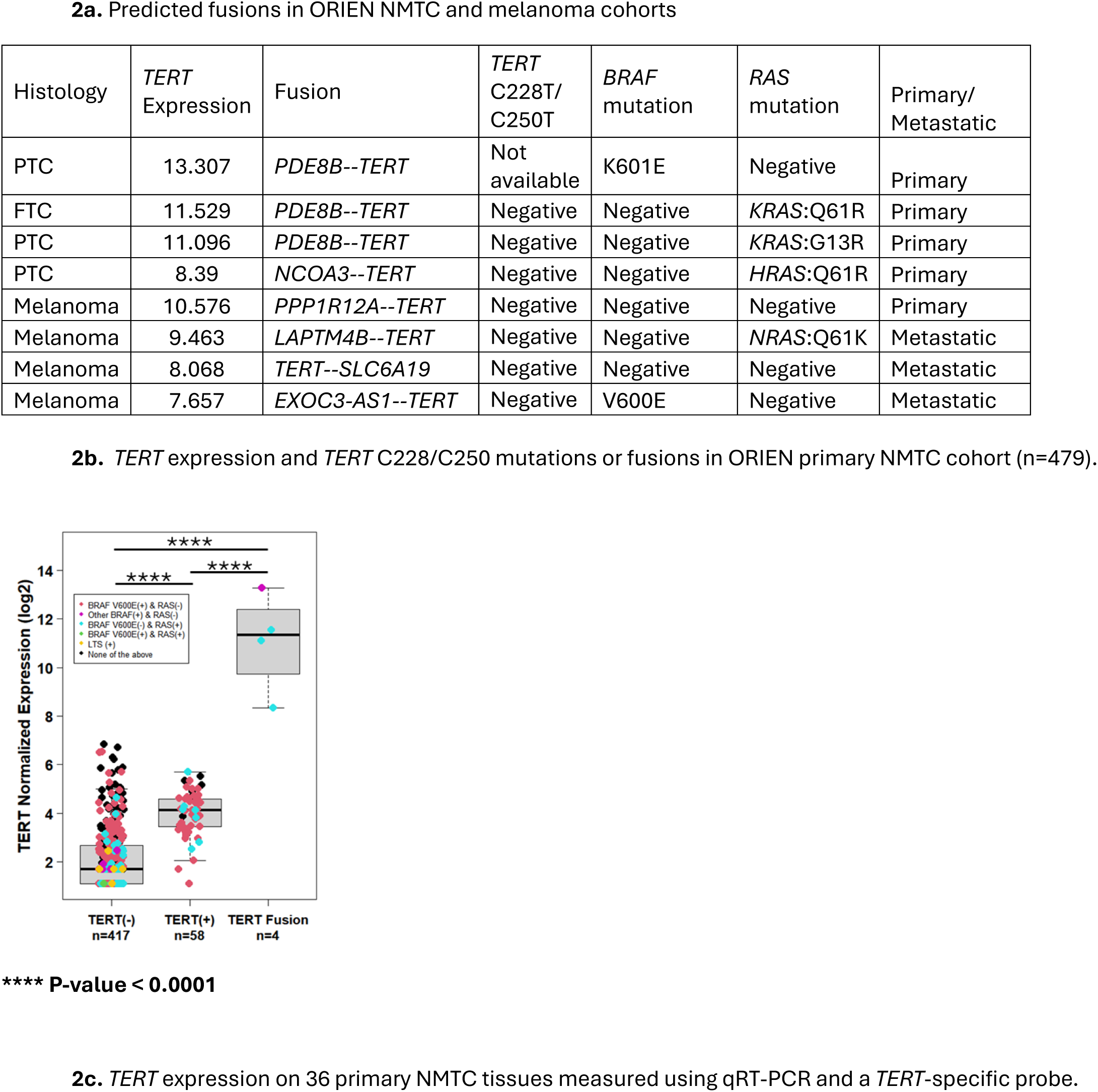
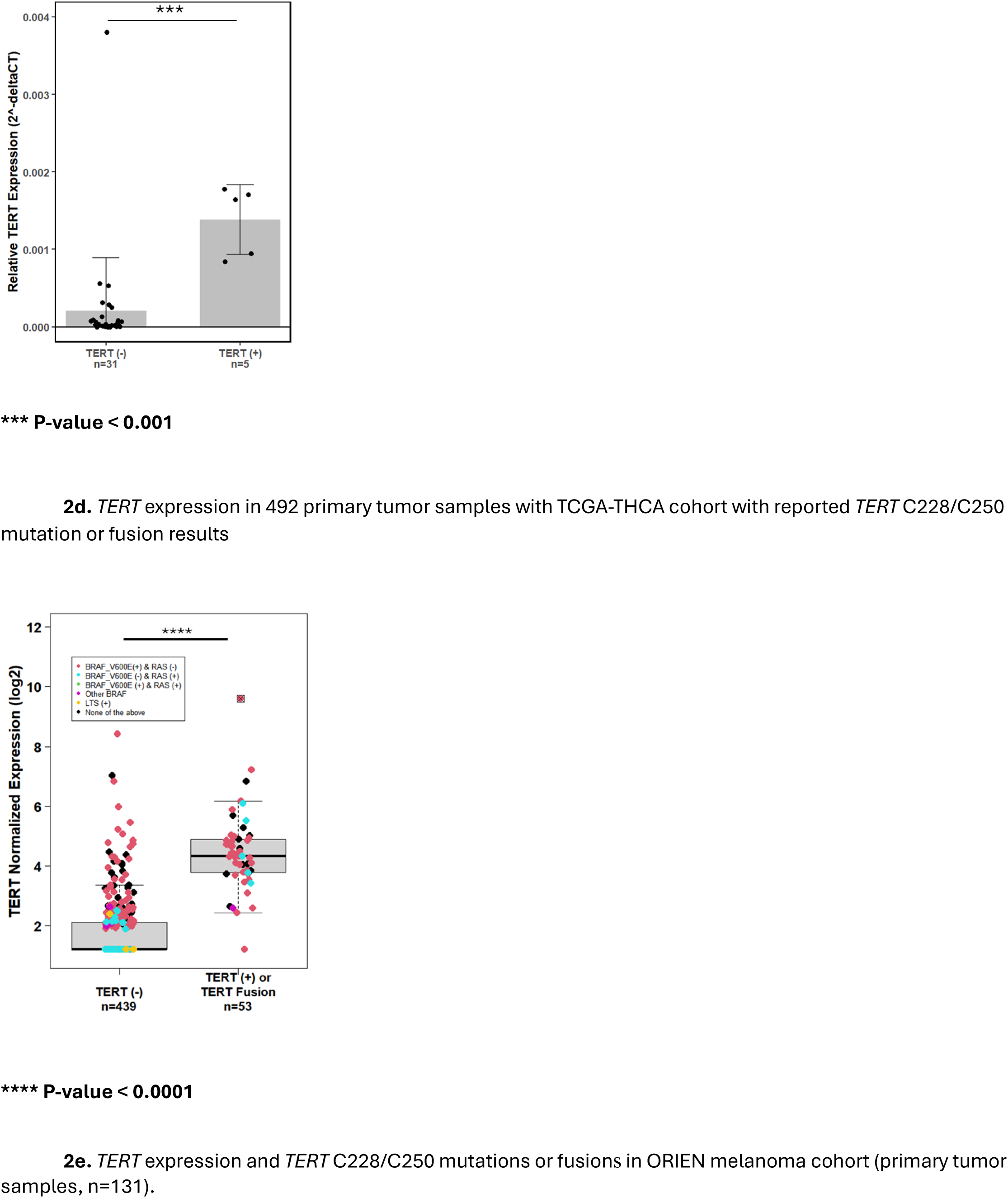
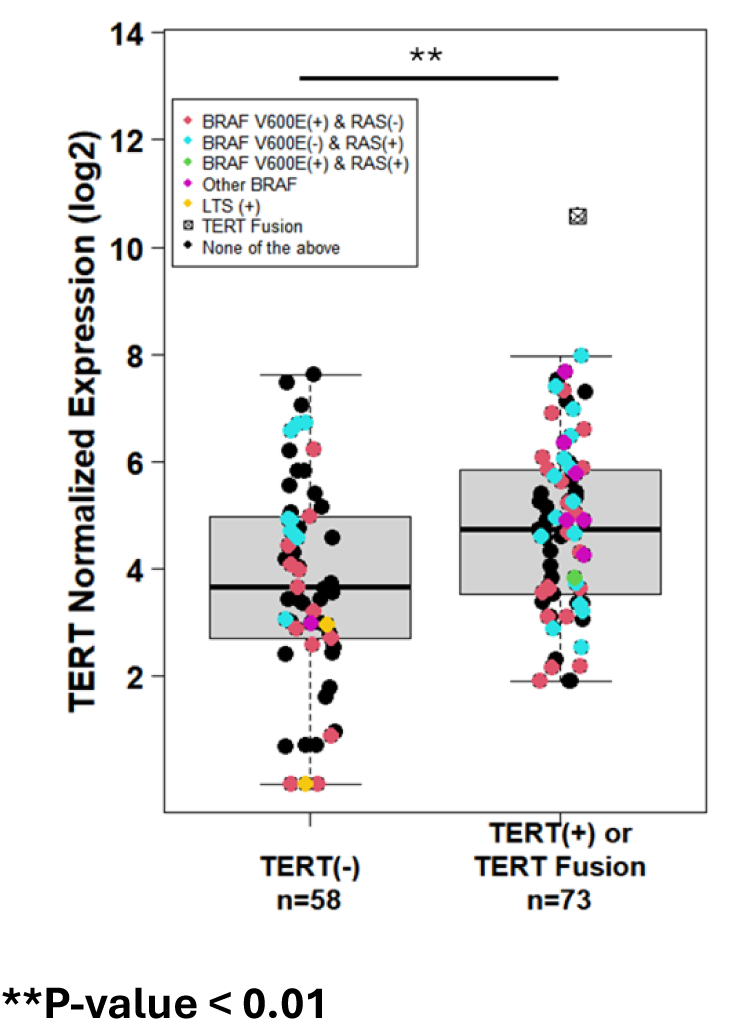
*TERT* expression and mutations in NMTC and melanoma. 2a. Predicted fusions in ORIEN NMTC and melanoma cohorts. 2b: *TERT* expression and *TERT* C228/C250 mutations or fusion status in ORIEN primary NMTC cohort (n=479). **** p<0.0001. 2c: *TERT* expression in 36 primary NMTC tissues measured using qRT-PCR and a *TERT*-specific probe. *** p<0.001. 2d. *TERT* expression in 492 primary tumor samples with TCGA-THCA cohort with reported *TERT* C228/C250 mutation or fusion results^28^. **** p<0.0001. 2e. Primary melanoma samples with relationship between *TERT* gene expression and *TERT* C228/C250 mutations or fusions in the ORIEN melanoma cohort (n=131). **p<0.01.

### *TERT* expression

Genomic alterations of 891 primary and 166 metastatic NMTC patients are shown in an Oncoplot (**Figure 1b**). Primary NMTC harboring *TERT* promoter mutations had higher *TERT* expression with average log2 fold increase of 1.83, (p<4e-11) while patients with *TERT* fusions had an average log2 fold increase of 8.92 (p<4e-11) (**Figure 2b** and **eTable 3a**). Metastatic NMTC showed a similar trend to primary tumors with average log2 fold increase of 0.674 (p=0.078) but without statistical significance (**eFigure 4a** & **eTable 4a**).

*TERT* expression also was assessed using TaqMan RT-qPCR in a separate cohort of 36 primary NMTC frozen tissue specimens available at Ohio State, including 5 samples with *TERT* promoter mutations. Samples with *TERT* promoter mutations showed increased *TERT* expression (**Figure 2c**, p=0.0001). A similar result was observed (p=0.002) between metastatic NMTC samples with (n=9) and without (n=12) *TERT* promoter mutations (**eFigure 4b**). Analysis in the 492 TCGA-THCA primary samples with *TERT* promoter mutations or fusion showed increased expression with log2 fold increase of 2.6 (p<2e-16) (**Figure 2d** & **eTable 3b**).

Because some of the somatic samples from ORIEN had uncertain tumor cell percentage, we performed a secondary analysis restricted to primary NMTC samples with tumor percentage ≥70% to align with TCGA. In this analysis, tumors with *TERT* promoter mutations had an average log2 fold increase of 1.93 (p<2e-16) and samples with fusions had an average log2 fold increase of 8.96 (p<2e-16; **eFigure 4c** & **eTable 4b**).

Primary NMTC samples with T4-stage had higher *TERT* expression versus smaller tumors (p < 0.05). There was no significant difference in *TERT* expression between T0-T3 stages (**Figure 3a**). Primary NMTC samples from patients with distant metastasis (M1) had significantly higher *TERT* expression compared to M0 tumors (p<0.0001) (**Figure 3a**). There was no significant difference in *TERT* expression related to nodal metastasis (**Figure 3a**). Similar relationships between *TERT* expression and staging were seen in the TCGA-THCA samples (**Figure 3b**).

**Figure 3.**
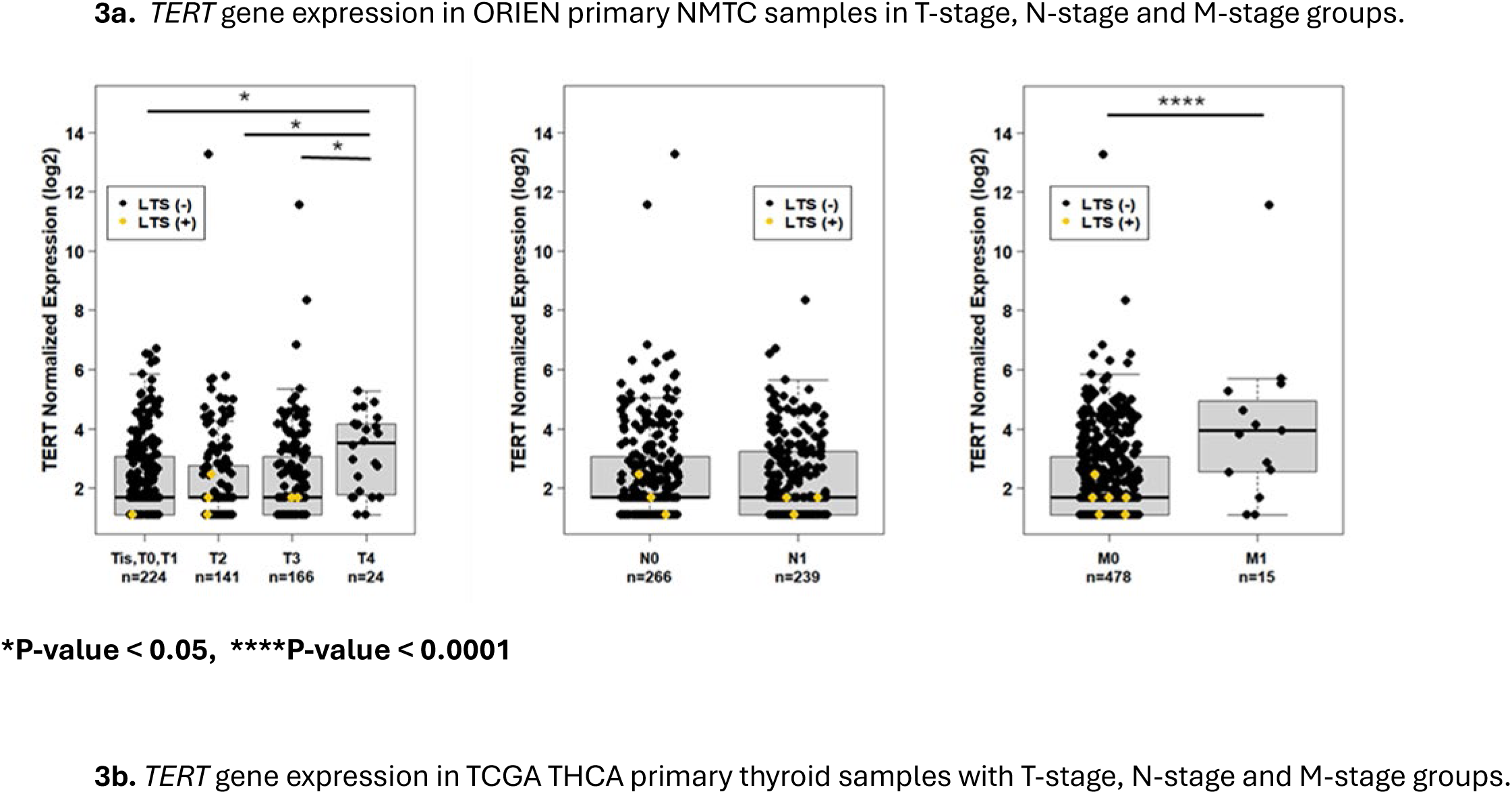
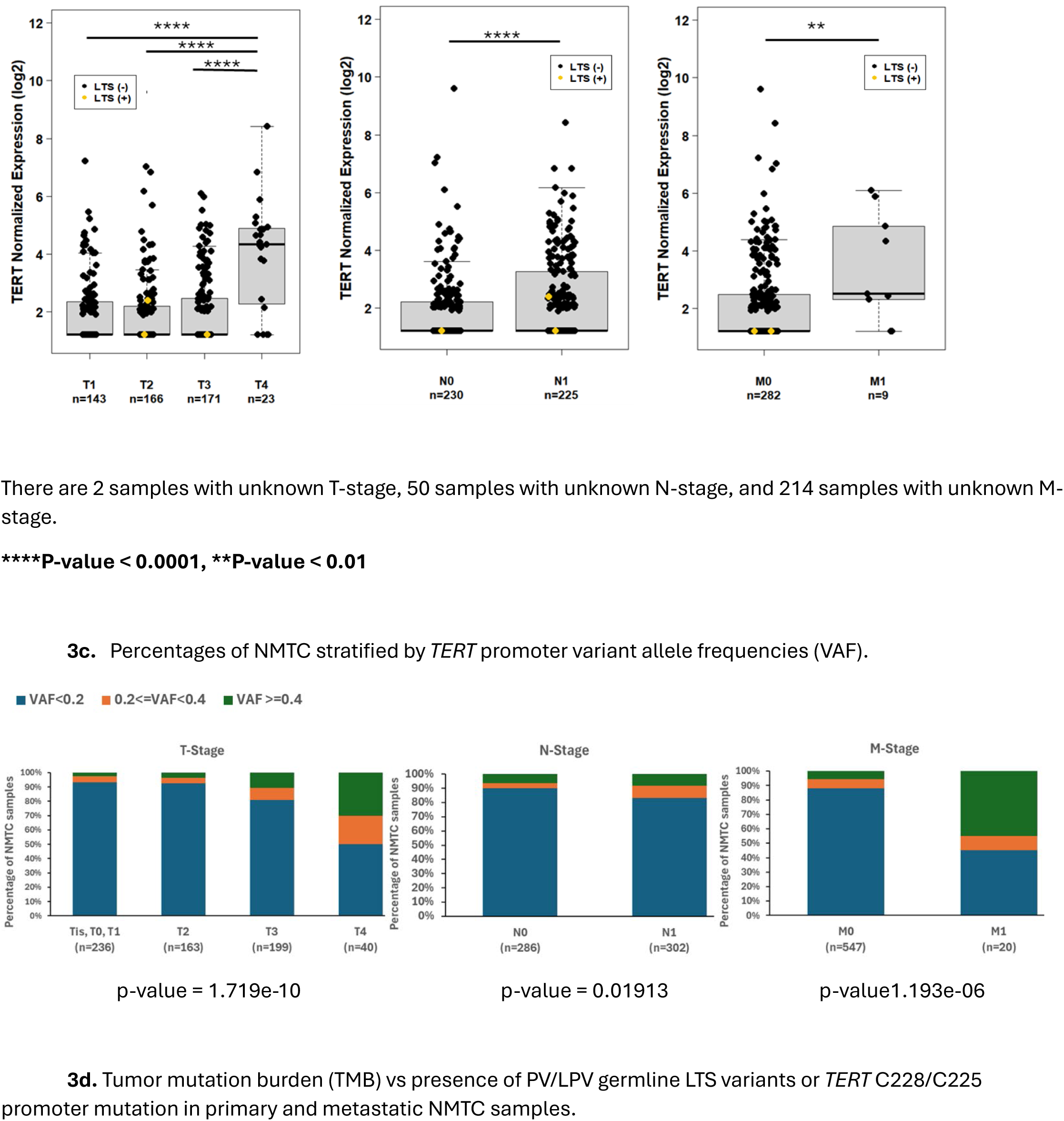
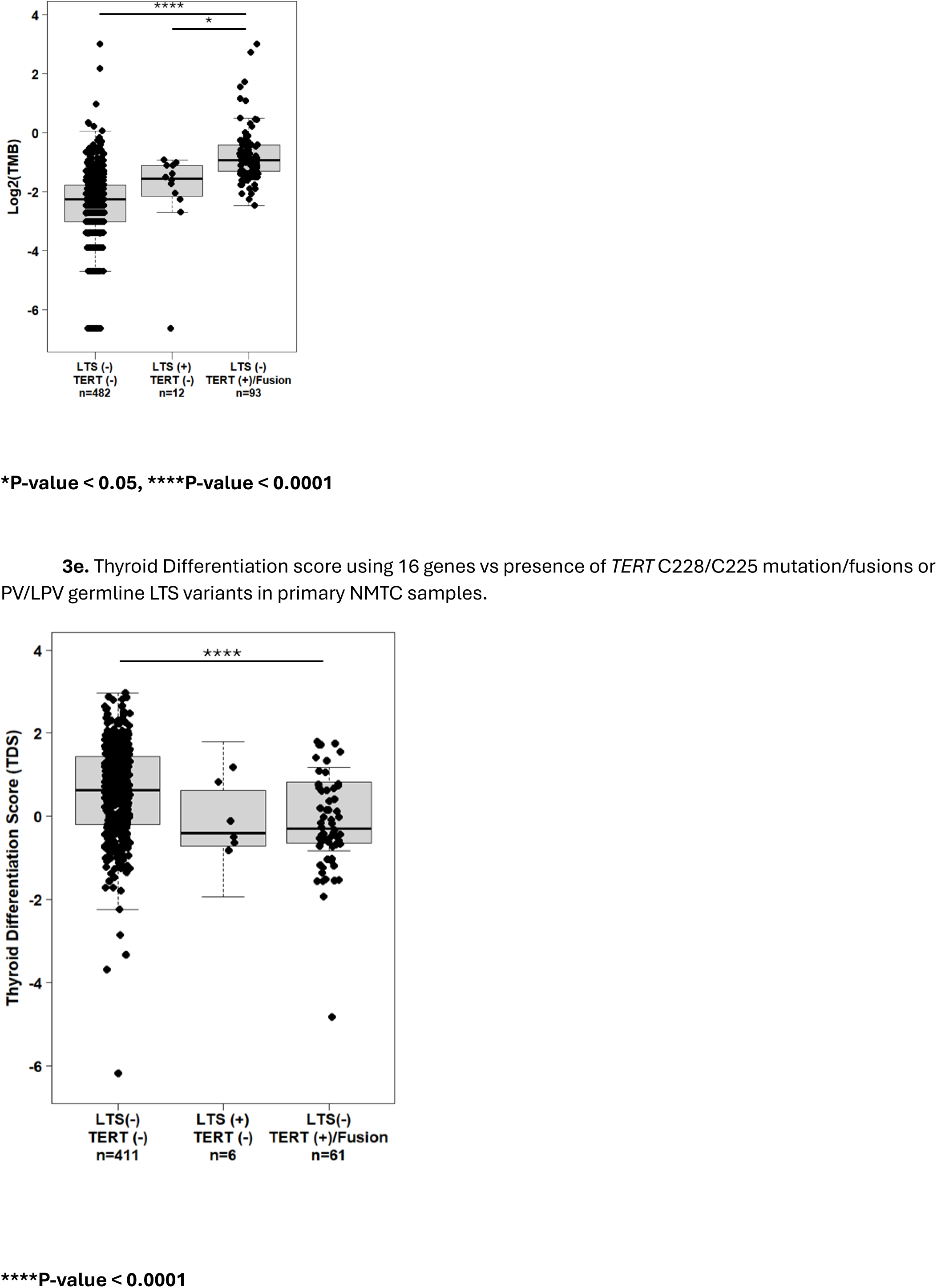
*TERT* expression in NMTC and melanoma, thyroid differentiation score and TMB. 3a: *TERT* gene expression in ORIEN primary NMTC samples in T-stage, N-stage and M-stage groups. There are 19 samples with unknown T-stage, 69 samples with unknown N-stage and 81 samples with unknown M-stage. *p<0.05, ****p<0. 0001. 3b: *TERT* gene expression in TCGA THCA primary thyroid samples with T-stage, N-stage and M-stage groups. There are 2 samples with unknown T-stage, 50 samples with unknown N-stage and 214 samples with unknown M-stage. ****p<0.0001, **p<0.01. 3c: Percentages of NMTC stratified by *TERT* promoter variant allele frequencies (VAF). T-stage p=1.719e-10; N-stage p=0.01913; M-stage p=1.193e-06. 3d: Tumor mutation burden (TMB) vs presence of PV/LPV germline LTS variants or *TERT* C228/C225 promoter mutation in primary and metastatic NMTC samples. *p<0.05, ****p<0.0001. 3e: Thyroid differentiation score using 16 genes vs presence of *TERT* promoter mutations or fusions and germline LTS variants in primary NMTC cohort.

Primary melanoma samples with *TERT* promoter mutations or *TERT* fusions had an average log2 fold increase of 1.019 (p=0.002) in *TERT* expression (**Figure 2e** & **eTable 3c**). Analysis of metastatic melanoma patients with and without *TERT* promoter mutations shows an average log2 fold increase of 1.027 (p=0.0002) (**eFigure 5** & **eTable 4c**).

There were 574 NMTC patients with germline WES and *TERT* expression data from the primary tumor available (**eFigure 6**); six had a LTS germline variant all of which had lower log2 *TERT* expression compared to patients without LTS variants, with a log2 fold change of −0.896 that was not statistically significant (95% CI=-2.08-0.29, p=0.14, **eTable 5**).

### *TERT* Promoter VAF, Tumor Mutation Burden, and Thyroid Differentiation

*TERT* promoter variant allele frequencies (VAF) were evaluated among NMTC tumors harboring a somatic variant. The proportion of patients in higher VAF groups increased as the severity of T-stage (p=1.719e-10), N-stage (p=0.019) or M-stage (p=1.193e-06) progressed (**Figure 3c**). We assessed the relationship between tumor mutation burden (TMB) and telomere-maintenance mechanism.

TMB among the NMTC patients with *TERT* promoter variant had an average log2 fold increase of 1.032 (p=0.02) versus patients with PV/LPV germline LTS variants (**Figure 3d** and **eTable 6**). There was no statistically significant difference in thyroid differentiation scores (TDS)^28^ between primary patients with *TERT* C228/C225 mutations or fusions and primary patients with germline LTS variants (**Figure 3e** & **eTable 7**). However, the presence of either telomere maintenance mechanism was associated with a lower TDS versus patients who did not (p=0.018).

## DISCUSSION

NMTC and melanoma, tumors commonly seen in inherited LTS, are often driven by somatic activating *BRAF* or *RAS* variants and have high frequencies of somatic *TERT* promoter mutations providing a framework for understanding germline/somatic telomere maintenance mechanisms. The current data reinforce the concept that inherited telomere lengthening variants appear mutually exclusive from somatic mechanisms, providing genetic evidence for overlapping roles in tumorigenesis. In the combined NMTC cohorts with both germline and somatic data, 0/20 patients with LTS had a somatic *TERT* promoter mutation or fusion, a statistically significant mutually exclusive relationship (p=0.0496) which also was identified on meta-analysis with the melanoma ORIEN cohort (p=0.036). In NMTC this relationship appears particularly relevant for PTC driven by *BRAF* or *RAS* mutations since they have been present in nearly all reported NMTC associated with a germline LTS variant. Further research is needed to define mechanisms by which cells that maintain telomere length avoid MAPK induced senescence; however, the genetic data support that telomere lengthening may be an early event in NMTC similar to melanoma^29–31^, which has also been suggested by a recent GWAS^32^.

In addition to germline LTS and somatic *TERT* alterations, alternative lengthening of telomeres (ALT) and other mechanisms by which telomere length is maintained may contribute to thyroid tumorigenesis, since they do not account for all NMTCs. While ALT is a common occurrence in cutaneous melanomas, it is reported to be rare in thyroid cancer^33^. Further studies of mechanisms for telomere maintenance are still needed in NMTC.

Although these germline and somatic changes appear to be mutually exclusive, the clinical associations are not overlapping in NMTC. The *TERT* promoter VAF data suggest that the initial somatic mutations may be subclonal but provide a growth advantage leading to progression. This is supported by the association between somatic *TERT* mutations and greater TMB. Moreover, tumors with somatic *TERT* promoter mutation/fusion were associated with higher levels of *TERT* expression, larger tumors, and metastasis while those from patients with germline LTS mutations were not. These associations may inform clinical care and suggest functional differences between these two telomere-lengthening mechanisms that require further research.

Consistent with previous studies, germline LTS LPV/PV were identified in 1.5% of unselected patients with NMTC and 0.5% of unselected patients with melanoma^1,34^. While telomere lengthening germline variants are uncommon in unselected patients with NMTC and melanoma, prior studies have shown enrichment in both familial thyroid cancer^1^ and melanoma^35–37^. Thus, it may be possible to identify at-risk individuals to inform genetic testing and counseling. RNA sequencing also identified *TERT* fusions associated with high levels of *TERT* expression, including previously reported and novel fusion partners^38^, highlighting the need for comprehensive tumor evaluation in clinical practice^11^.

In summary, we provide evidence that germline LTS variants and somatic *TERT* promoter mutations are mutually exclusive and are enriched for *BRAF*/*RAS*-driven PTC suggesting that telomere lengthening events are critical for MAPK-driven thyroid tumorigenesis. However, aggressive clinical features and genomic instability are specifically associated with somatic *TERT* promoter variants suggesting differential effect on cancer biology. These results have mechanistic relevance and can inform clinical care for patients at risk of or that have thyroid cancer.

## Author contributions

SL: Data Analysis; Data Review, Manuscript Writing, Review and Approval

PLB: Conceptualization, Patient Enrollment, Data Analysis; Data Review, Manuscript Writing, Review and Approval

TTN: Data Review, Manuscript Writing, Review and Approval

WL: Research Experiments; Data Review, Data Analysis; Data Review, Manuscript Writing, Review and Approval

NP, BRH, HM, BS, KJ, ARN, VK, and JF: Patient Enrollment, Data Review, Manuscript Writing, Review and Approval

MDR: Conceptualization, Patient Enrollment, Data Analysis; Research Experiments; Data Review; Manuscript Writing, Review, and Approval

## Acknowledgements

We would like to acknowledge Mary Armanios, MD (Johns Hopkins, Baltimore Maryland) for critical review of data and manuscript review. We also acknowledge Kathleen Egan, ScD (Moffitt Cancer Center, Tampa FL), Therese Bocklage, MD (University of Kentucky Markey Cancer Center, Lexington, KY), Amanda M. Laird, MD (Rutger Robert Wood Johnason Medical School and Cancer Institute, New Brunswick, NJ), and Stephen Edge, MD (Roswell Park Comprehensive Cancer Center, Buffalo, NY) for patient enrollment in ORIEN.

## Data Availability Statement

Data from the Oncology Research Information Exchange Network (ORIEN) is accessible through project requests.

