## Supplemental figures and tables for "Telomere maintaining germline and somatic variants in thyroid cancer and melanoma"

**Online-Only Materials**

**eFigure 1.** *TERT* gene and promoter region coverage by capture kits used in the WES analysis from the UCSC genome browser.

**eMethods.**

**eFigure 2.** NMTC cohort distribution with respect to *BRAF/RAS* variant status, *TERT* promoter status, and availability of *TERT* expression data.

**eFigure 3.** Melanoma cohort distribution with respect to *BRAF/RAS* variant status, *TERT* promoter status, and availability of *TERT* expression data.

**eTable 1.** LPV/PV in *ACD, POT1* and *TINF2* from ORIEN NMTC, DeBoy et al[1], TCGA-THCA, and ORIEN melanoma cohorts.

**eTable 2.** Association between *TERT* promoter mutation status and T/M stages in the ORIEN NMTC cohort.

**eTable 3.** Univariate and multivariable analysis of ORIEN primary NMTC, TCGA-THCA, ORIEN primary melanoma comparing *TERT* expression between *TERT* C228/C250 mutations or fusion status.

**eFigure 4.** Relationships between TERT expression and TERT promoter mutation status in ORIEN metastatic NMTC, OSU metastatic NMTC, and ORIEN NMTC with >70% tumor content.

**eTable 4.** Univariate and multivariable analysis of ORIEN metastatic NMTC, primary NMTC with >70% tumor content, and metastatic melanoma.

**eFigure 5.** Metastatic melanoma with *TERT* gene expression and known *TERT* promoter mutation status.

**eFigure 6.** Primary NMTC samples with *TERT* gene expression and known germline *ACD*, *POT1*, and *TINF2* PV/LPV status.

**eTable 5.** Univariate and multivariable analysis results of 574 primary NMTC patients comparing *TERT* expression by germline *ACD*, *POT1*, and *TINF2* PV/LPV status.

**eTable 6.** Univariate and multivariable analysis results comparing log2(TMB) (Tumor mutation burden) vs presence of PV/LPV germline LTS variants and *TERT* C228/C225 mutation in primary and metastatic NMTC samples

**eTable 7.** Thyroid differentiation score (TDS) using 16 genes vs presence of *TERT* C228/C225 mutation/Fusions or PV/LPV germline LTS variants in primary NMTC samples

**Online-Only References**

**eFigure 1.** *TERT* gene and promoter region coverage by capture kits used in the WES analysis from the UCSC genome browser.

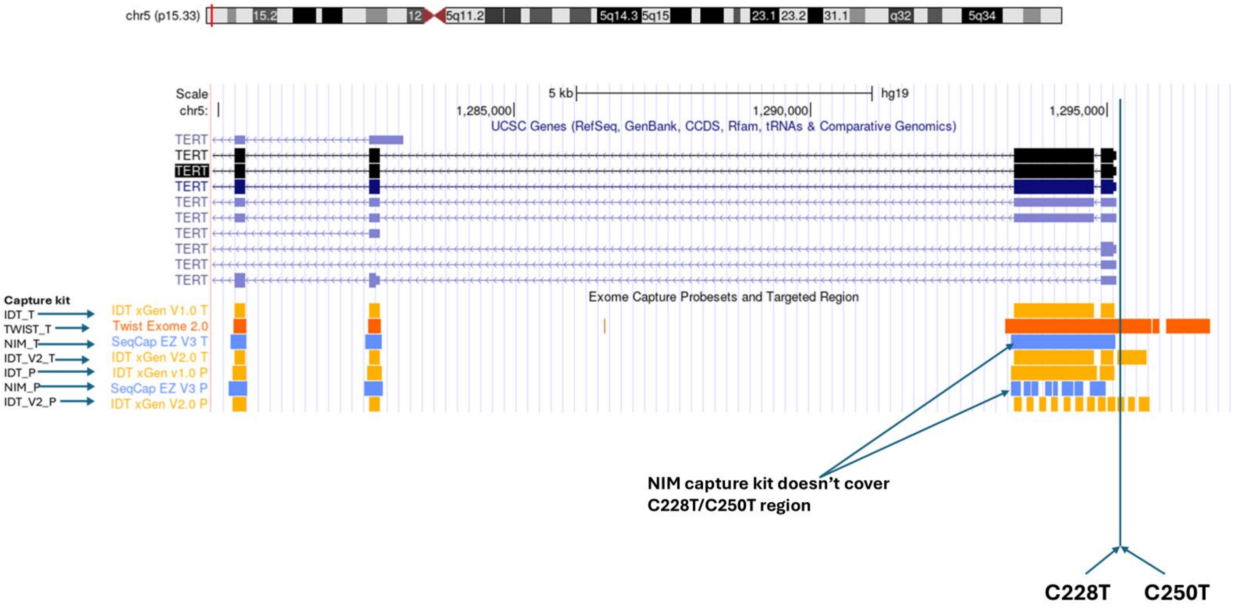

### eMethods

#### Detailed Statistical Analysis:

Linear models were applied to compare *TERT* expression between groups adjusting for covariates (age of diagnosis and sex). Variation inflation factors were estimated to evaluate the severeness of multi[2]collinearity in multivariable models. To confirm that % tumor content (%TC) has no bias on the results, similar analysis using patient samples with %TC>=70 was performed. TaqMan *TERT* expression data were compared by applying non-parametric Wilcoxon Rank Sum tests. Fisher's exact test was used for categorical data, with Lancaster mid-P method[2] to test associations with zero count cells. Odds ratios and their 95% confidence intervals were estimated with Haldane-Anscombe correction. R libraries 'exact2x2' and 'epitools' were used for the analysis. Tumor mutation burden (TMB) estimates were compared between germline LTS/*TERT* promoter mutation groups by applying linear models adjusting for %TC, age of diagnosis, and sex. Arcsine sqrt transformation was applied to %TC in the model. Thyroid differentiation score (TDS)[3] was estimated using 16 genes and compared between germline LTS/*TERT* promoter mutation groups by applying linear models. Tukey's post-hoc tests were applied to assess the significance of differences between pairs of group means when appropriate. Cochran-Mantel-Haenszel chi-squared test was applied to get the meta-analysis of thyroid and melanoma studies.

**eFigure 2.** NMTC cohort distribution with respect to *BRAF/RAS* variant status, *TERT* promoter status, and availability of *TERT* expression data.

**eFigure 2a.** NMTC cohort distribution (primary tumors)

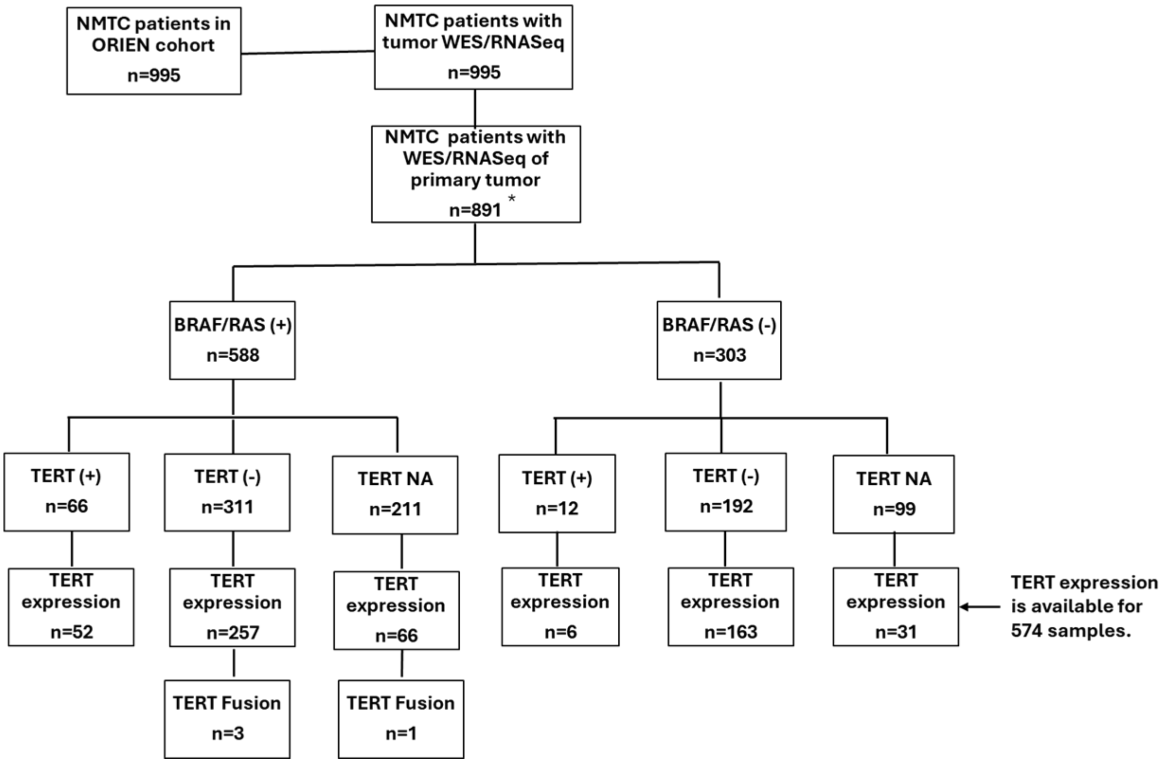

* 62 patients had both primary and metastatic tumors.

**eFigure 2b.** NMTC cohort distribution (metastatic tissues).

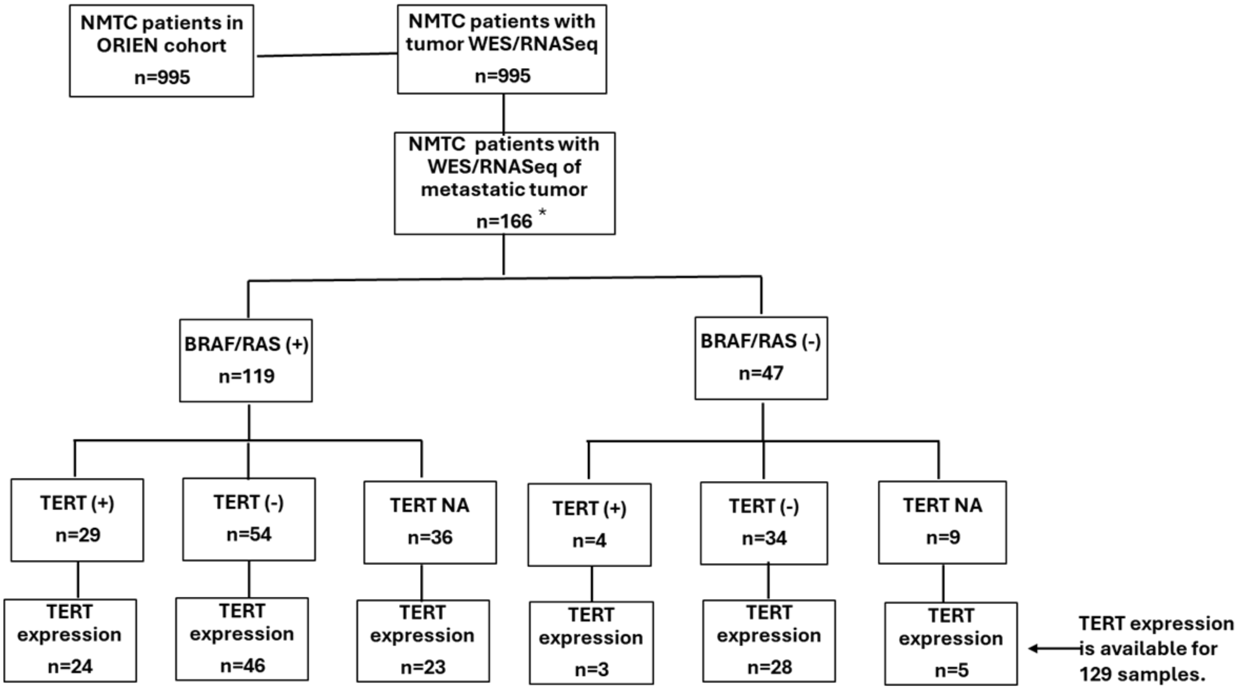

* 62 patients had both primary and metastatic tumors.

**eFigure 3.** Melanoma cohort distribution with respect to *BRAF/RAS* variant status, *TERT* promoter status, and availability of *TERT* expression data.

**eFigure 3a.** Melanoma cohort distribution (primary tumors). The melanoma cohort included 400/993 patients with germline sequencing data and 593/993 patients with both tumor and germline sequencing data.

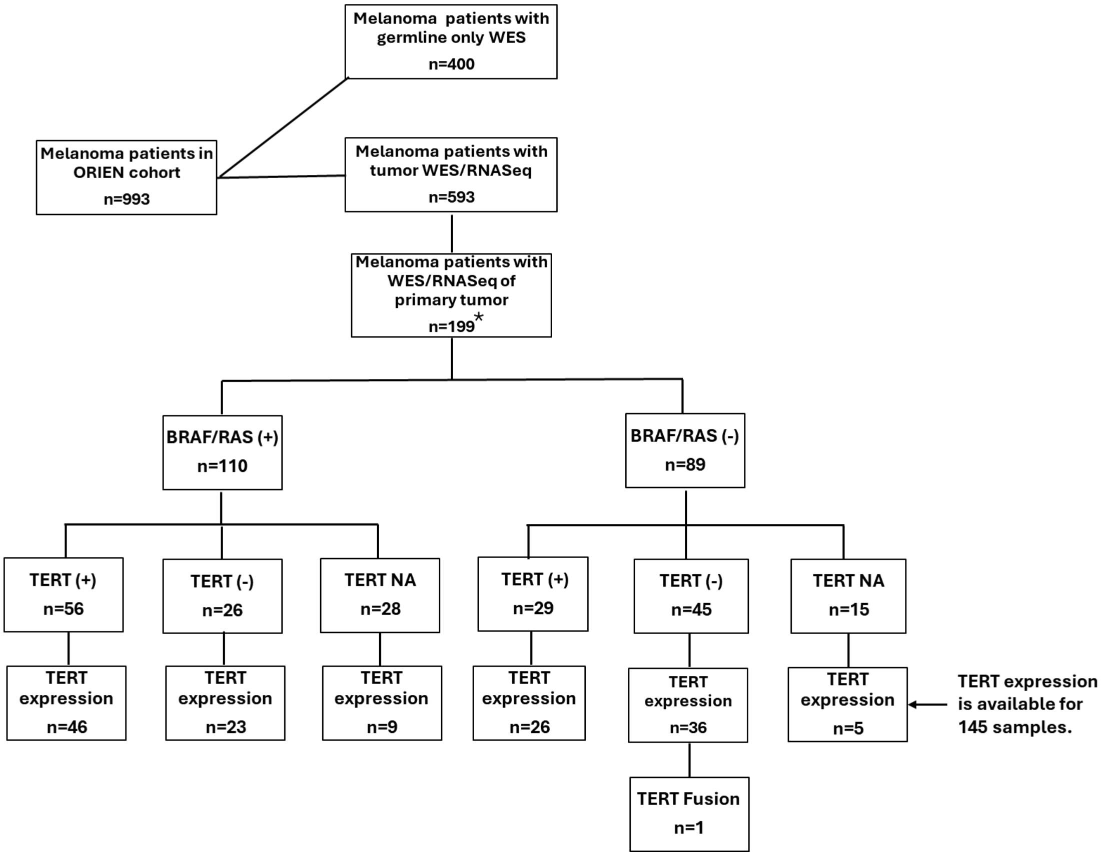

*21 patients with both primary and metastatic tumors sequenced.

**eFigure 3b**. Melanoma cohort distribution (metastatic tumors).

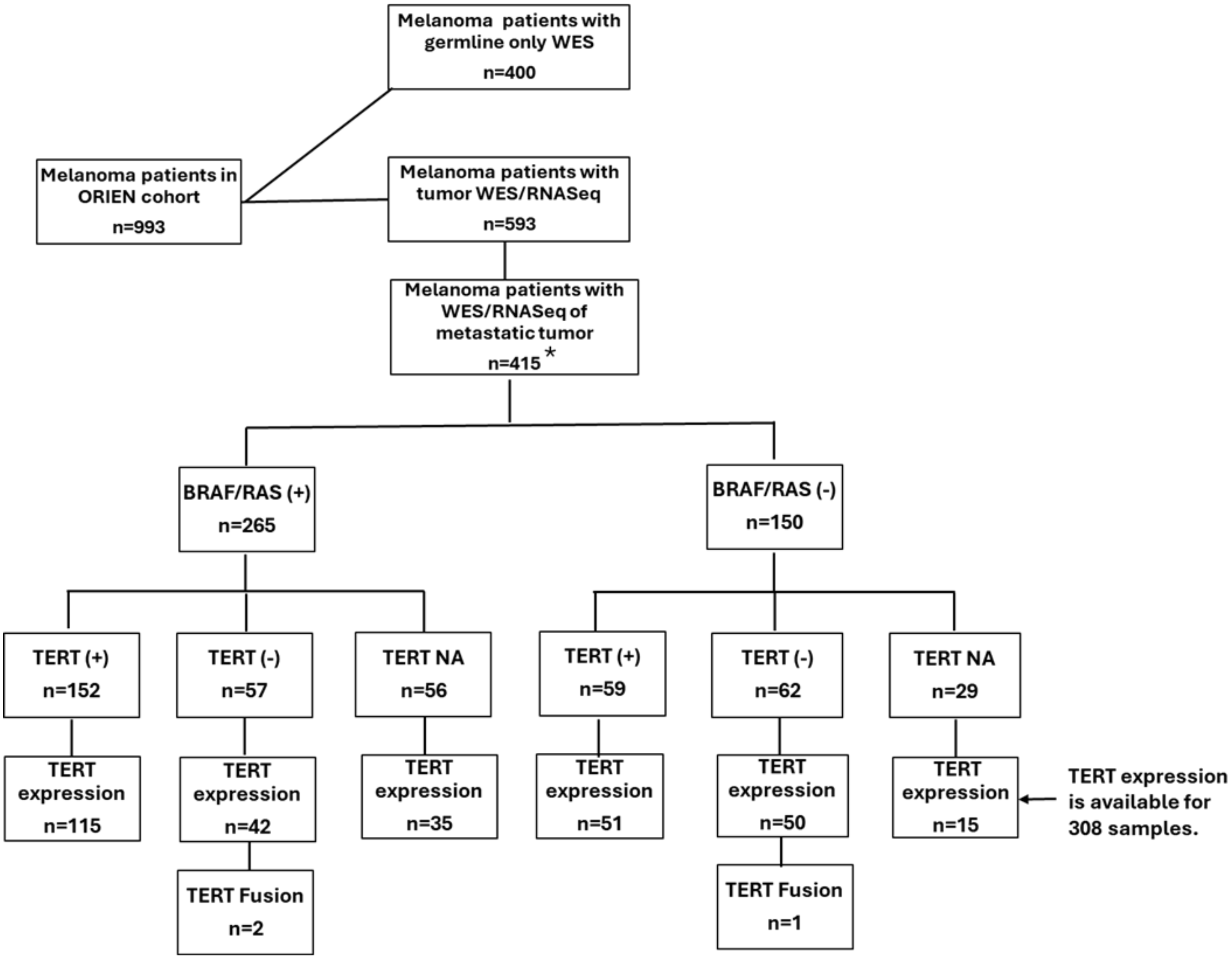

*21 patients with both primary and metastatic tumors sequenced.

**eTable 1. LPV/PV in *ACD, POT1* and *TINF2* from ORIEN NMTC, DeBoy et al[1], TCGA-THCA, and ORIEN melanoma cohorts. Genomic positions are in hg38.**

| **Diagnosis** | **Gene** | Variant location | **Coding variant** | **Protein impact** | ***TERT* C228T/ C250T** | ***BRAF* V600E** | ***RAS* LPV/PV** | **T** | **N** | **M** | **Cohort** |
|---|---|---|---|---|---|---|---|---|---|---|---|
| PTC | *ACD* | 16:67660121:C:T | c.99+1G>A | - | negative | positive | negative | T2 | N0 | M0 | TCGA |
| PTC | *ACD* | 16:67659536:C:G | c.413+1G>C | - | negative | positive | negative | T1 | N0 | M0 | ORIEN/ DeBoy et al |
| PTC | *ACD* | 16:67659234:T:C | c.488A>G | p.Asn163Ser | NA | positive | negative | T3 | N1 | M0 | ORIEN |
| PTC | *ACD* | 16:67659234:T:C | c.488A>G | p.Asn163Ser | negative | positive | negative | T1 | N1 | MX | ORIEN/ DeBoy et al |
| PTC | *ACD* | 16:67659234:T:C | c.488A>G | p.Asn163Ser | negative | negative | negative | T2 | N1 | MX | TCGA |
| PTC | *POT1* | 7:124892268:T:C | c.121A>G | p.Thr41Ala | negative | positive | NA | T3 | N1 | M0 | DeBoy et al |
| PTC | *POT1* | 7:124871018:AA:A | c.147del | p.Ile49Metfs*7 | negative | positive | negative | T2 | N1 | M0 | ORIEN |
| PTC | *POT1* | 7:124870933:A:G | c.233T>C | p.Ile78Thr | negative | negative | NRAS:Q61R | T3 | N0 | M0 | ORIEN |
| PTC | *POT1* | 7:124870913:T:C | c.253A>G | p.Lys85Glu | negative | positive | negative | T1 | N1 | M0 | ORIEN/ DeBoy et al |
| PTC | *POT1* | 7:124853035:G:C | c.806C>G | p.Thr269Ser | negative | positive | negative | T2 | N1 | M0 | ORIEN |
| PTC | *POT1* | 7:124842898:A:AA | c.1071dup | p.Gln358Serfs*13 | negative | positive | NA | T3 | N1 | M0 | DeBoy et al |
| PTC | *POT1* | 7:124842898:A:AA | c.1071dup | p.Gln358Serfs*13 | negative | positive | NA | T2 | N0 | M0 | DeBoy et al |
| PTC | *POT1* | 7:124841179:C:T | c.1164-1G>A | - | negative | negative * | negative | T1 | N1 | M0 | ORIEN |
| PTC | *POT1* | 7:124835303:A:T | c.1481T>A | p.Ile494Lys | negative | positive | negative | T3 | N1 | M0 | ORIEN/ DeBoy et al |
| PTC | *POT1* | 7:124825277:CAT:C | c.1765_1766del | p.Met589Valfs*9 | negative | positive | negative | T3 | N1 | M0 | TCGA |
| PTC | *TINF2* | 14:24242332:T:C | c.1A>G | p.Met1? | negative | positive | negative | T3 | N1 | M0 | ORIEN/ DeBoy et al |
| PTC | *TINF2* | 14:24241937:CA: | c.248_249del | p.Leu83Glnfs*53 | negative | positive | NA | T1 | N1 | M0 | DeBoy et al |
| PTC | *TINF2* | 14:24241675:C:G | c.399G>C | p.Gln133His | negative | positive | negative | T2 | N0 | M0 | ORIEN |
| PTC | *TINF2* | 14:24241032:CC:C | c.591del | p.Trp198Glyfs*12 | negative | positive | NA | T1 | NX | MX | DeBoy et al |
| PTC | *TINF2* | 14:24240686:G:A | c.793C>T | p.Arg265* | negative | positive | NA | T1 | N1 | M0 | DeBoy et al |
| PDTC | *TINF2* | 14:24240687:G:A | c.793C>T | p.Arg265* | negative | negative | KRAS:G12V | T1 | N0 | M0 | ORIEN |
| **Diagnosis** | **Gene** | Variant location | **Coding variant** | **Protein impact** | ***TERT* C228T/ C250T** | ***BRAF* V600E** | ***RAS* LPV/PV** | **T** | **N** | **M** | **Cohort** |
| Melanoma | *ACD* | 16:67659252:G:C | c.470C>G | p.Ser157Ter | NA | NA | NA | NA | NA | NA | ORIEN |
| Melanoma | *POT1* | 7:124870933:A:G | c.233T>C | p.Ile78Thr | NA | NA | NA | T0 | N0 | M0 | ORIEN |
| Melanoma | *POT1* | 7:124870913:T:C | c.253A>G | p.Lys85Glu | NA | NA | NA | NA | NA | NA | ORIEN |
| Melanoma | *POT1* | 7:124858983:G:T | c.676C>A | p.His226Asn | negative | negative | No | T3 | NA | M0 | ORIEN |
| Melanoma | *POT1* | 7:124842858:G:A | c.1112C>T | p.Pro371Leu | negative | negative | NRAS:Q61K | T4 | N0 | M0 | ORIEN |
| Melanoma | *POT1* | 7:124835276:T:C | c.1505+3A>G | - | NA | NA | NA | NA | NA | NA | ORIEN |
| Melanoma | *POT1* | 7:124824065:G:A | c.1802C>T | p.Pro601Leu | NA | NA | NA | NA | NA | NA | ORIEN |

* *BRAF* V600E VAF < 5%. PTC = papillary thyroid cancer. PDTC = poorly differentiated thyroid cancer. NA = Not assessed.

**eTable 2.** Association between *TERT* promoter mutation status and T/M stages in the ORIEN NMTC cohort.

| | *TERT* C228/C250 mutations (+) or Fusions | *TERT* C228/C250 mutations (-) |
|---|---|---|
| T4 or M1 | 31 | 22 |
| Tis, T0-T3 and M0 | 62 | 434 |

p-value = 3.04e-13, OR=9.80, 95%CI=(5.13, 19.00)

**eTable 3.** Univariate and multivariable analysis of ORIEN primary NMTC, TCGA-THCA, ORIEN primary melanoma comparing *TERT* expression between *TERT* C228/C250 mutations or fusion status.

**eTable 3a.** Univariate and multivariable analysis results of 479 ORIEN primary NMTC patients comparing *TERT* expression between *TERT* C228/C250 mutations or fusion status.

| | Univariate Analysis | | Multivariable Analysis | | |
|---|---|---|---|---|---|
| Variable | Log2FoldChange | P-value | Log2FoldChange | P-value | P-value with Post hoc Tukey's adjustment |
| Age at diagnosis | 0.014 | 0.001 | -0.000568 | 0.875 | |
| Male vs Female | 0.184 | 0.209 | -0.0361 | 0.756 | |
| *TERT*(+) vs *TERT* (-) | 1.816 | <2e-16 | 1.832 | <2e-16 | <4e-11 |
| *TERT* Fusion vs *TERT*(-) | 8.902 | <2e-16 | 8.918 | <2e-16 | <4e-11 |
| *TERT* Fusion vs *TERT*(+) | 7.086 | <2e-16 | 7.087 | <2e-16 | <4e-11 |

**eTable 3b.** Univariate and multivariable analysis results of 492 TCGA-THCA primary patients comparing *TERT* expression between *TERT* C228/C250 promoter mutations status.

| | Univariate Analysis | | Multivariable Analysis | |
|---|---|---|---|---|
| Variable | Log2FoldChange | P-value | Log2FoldChange | P-value |
| Age at diagnosis | 0.023 | 2.52E-10 | 0.004 | 0.213 |
| Male vs Female | 0.289 | 0.03 | 0.011 | 0.916 |
| *TERT*(+)/Fusion vs TERT(-) | 2.68 | <2e-16 | 2.6 | <2e-16 |

**eTable 3c.** Multivariable analysis results of 131 ORIEN primary melanoma patients comparing *TERT* expression between *TERT* C228/C250 promoter mutations or fusion status.

| | Univariate Analysis | | Multivariable Analysis | |
|---|---|---|---|---|
| Variable | Log2FoldChange | P-value | Log2FoldChange | P-value |
| Age at diagnosis | 0.001 | 0.927 | -0.003 | 0.8 |
| Male vs Female | 0.365 | 0.267 | 0.169 | 0.603 |
| *TERT*(+) or *TERT* Fusion vs *TERT*(-) | 1.044 | 0.001 | 1.019 | 0.002 |

**eFigure 4. Relationships between TERT expression and TERT promoter mutation status in ORIEN metastatic NMTC, OSU metastatic NMTC, and ORIEN primary NMTC with >70% tumor content.**

**eFigure 4a.** ORIEN metastatic NMTC patient samples with *TERT* expression and *TERT* promoter mutation status.

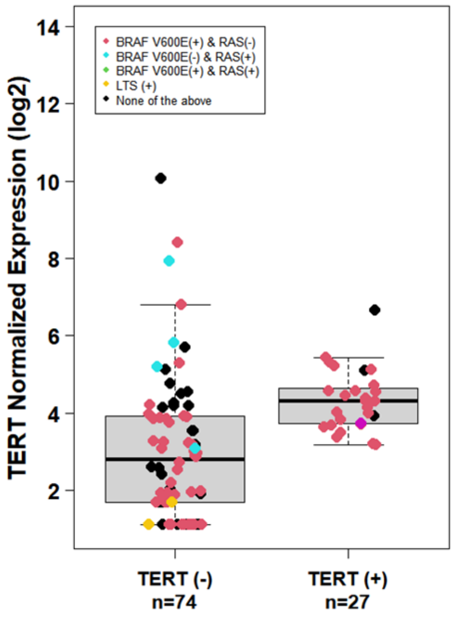

**P-value=0.078**

**eFigure 4b**. *TERT* expression in 21 OSU metastatic NMTC tissues measured by TaqMan assay

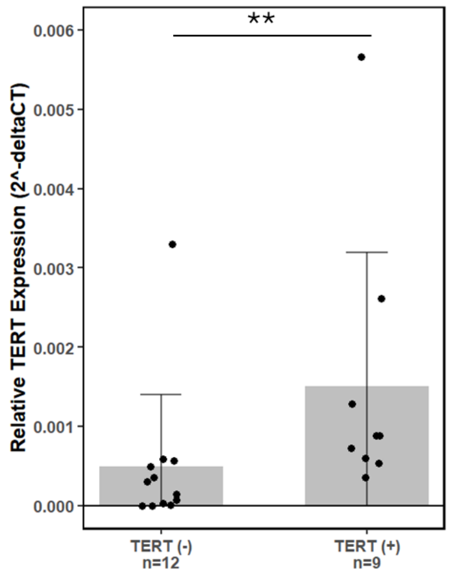

****P-value < 0.01**

Measured using qRT-PCR and a *TERT*-specific probe

**eFigure 4c.** There are 402 Primary NMTC patient samples with *TERT* gene expression and known *TERT* promoter mutation or fusion status that have %tumor content >=70.

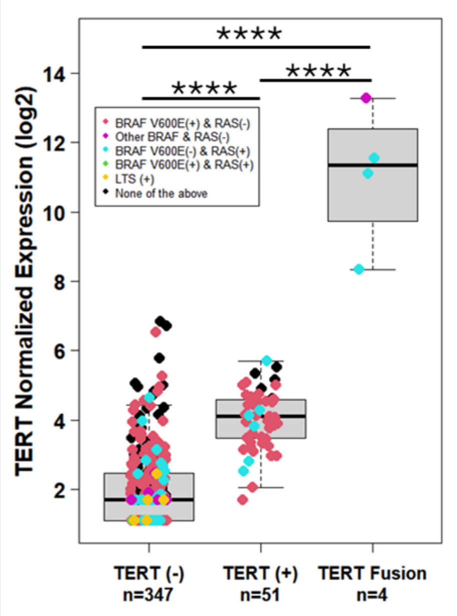

****** P-value < 0.0001**

**eTable 4.** Univariate and multivariable analysis of ORIEN metastatic NMTC, primary NMTC with >70% tumor content, and metastatic melanoma.

**eTable 4a.** Univariate and multivariable analysis results of 101 metastatic NMTC patients comparing *TERT* expression.

| | Univariate Analysis | | Multivariable Analysis | |
|---|---|---|---|---|
| Variable | Log2FoldChange | P-value | Log2FoldChange | P-value |
| Age at diagnosis | 0.044 | 1.23E-05 | 0.032 | 0.002 |
| Male vs Female | 0.992 | 0.004 | 0.587 | 0.07 |
| *TERT*(+) vs *TERT*(-) | 1.29 | 0.001 | 0.674 | 0.078 |

**eTable 4b.** Univariate and multivariable analysis results of 402 primary NMTC patients with known *TERT* promoter mutation or fusion status with %tumor content >=70, comparing *TERT* expression.

| | Univariate Analysis | | Multivariable Analysis | | |
|---|---|---|---|---|---|
| Variable | Log2FoldChange | P-value | Log2FoldChange | P-value | P-value with Post hoc Tukey's adjustment |
| Age at diagnosis | 0.021 | 5.73E-06 | 0.00317 | 0.36 | |
| Male vs Female | 0.303 | 0.051 | 0.0221 | 0.842 | |
| *TERT*(+) vs *TERT*(-) | 1.977 | <2e-16 | 1.926 | <2e-16 | <2e-16 |
| *TERT* Fusion vs *TERT*(-) | 9.026 | <2e-16 | 8.964 | <2e-16 | <2e-16 |
| *TERT* Fusion vs *TERT*(+) | 7.05 | <2e-16 | 7.04 | <2e-16 | <2e-16 |

**eTable 4c.** Univariate and multivariable analysis results of 258 metastatic melanoma patients comparing *TERT* expression.

| | Univariate Analysis | | Multivariable Analysis | |
|---|---|---|---|---|
| Variable | Log2FoldChange | P-value | Log2FoldChange | P-value |
| Age at diagnosis | 0.00756 | 0.397 | 0.006004 | 0.495 |
| Male vs Female | -0.0276 | 0.922 | -0.1292 | 0.64 |
| *TERT*(+) or *TERT* Fusion vs *TERT*(-) | 1.0291 | 0.00018 | 1.0269 | 0.0002 |

**eFigure 5.** Metastatic melanoma with *TERT* gene expression and known *TERT* promoter mutation status.

Three patients have *TERT* fusions. No samples with PV/LPV germline LTS variants.

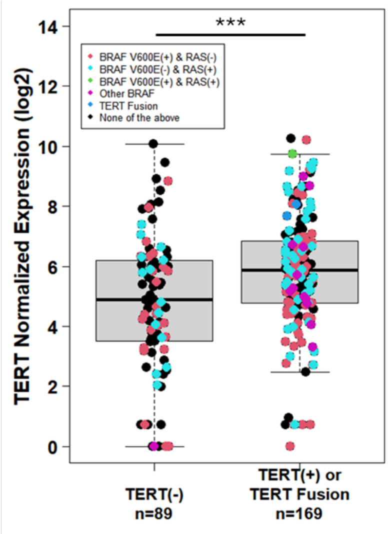

***** P-value < 0.001**

**eFigure 6.** Primary NMTC samples with *TERT* gene expression and known germline *ACD*, *POT1*, and *TINF2* PV/LPV status.

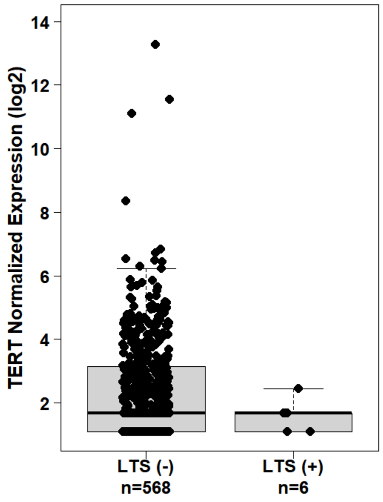

**P-value=0.159**

**eTable 5.** Univariate and multivariable analysis results of 574 primary NMTC patients comparing *TERT* expression by germline *ACD*, *POT1*, and *TINF2* PV/LPV status.

| | Univariate Analysis | | Multivariable Analysis | |
|---|---|---|---|---|
| Variable | Log2FoldChange | P-value | Log2FoldChange | P-value |
| Age at diagnosis | 0.016 | 3.66E-05 | 0.015 | 0.0002 |
| Male vs Female | 0.279 | 0.032 | 0.165 | 0.215 |
| LTS(+) vs LTS(-) | -0.794 | 0.195 | -0.896 | 0.139 |

**eTable 6.** Univariate and multivariable analysis results comparing log2(TMB) (Tumor mutation burden) vs presence of PV/LPV germline LTS variants and *TERT* C228/C225 mutation in primary and metastatic NMTC samples

| | Univariate Analysis | | Multivariable Analysis | | |
|---|---|---|---|---|---|
| Variable | Log2FoldChange | P-value | Log2FoldChange | P-value | P-value with Post hoc Tukey's adjustment |
| Age of Diagnosis | 0.032 | < 2e-16 | 0.022 | 1.56E-10 | |
| Male vs Female | 0.253 | 0.035 | -0.151 | 0.16 | |
| Tumor Content | 1.071 | 8.59E-07 | 0.9 | 2.33E-06 | |
| LTS(+) & *TERT*(-) vs LTS (-) & *TERT*(-) | 0.477 | 0.199 | 0.327 | 0.374 | 0.647 |
| LTS (-) & *TERT*(+)/Fusion vs LTS(+)&*TERT*(-) | 1.216 | 0.002 | 1.032 | 0.008 | 0.02 |
| LTS(-) & *TERT*(+)/Fusion vs LTS (-) & *TERT*(-) | 1.693 | < 2e-16 | 1.359 | < 2e-16 | < 6e-10 |

**eTable 7.** Thyroid differentiation score (TDS) using 16 genes vs presence of *TERT* C228/C225 mutation/Fusions or PV/LPV germline LTS variants in primary NMTC samples

| | Univariate Analysis | | Multivariable Analysis | | |
|---|---|---|---|---|---|
| Variable | Mean Change | P-value | Mean Change | P-value | P-value with Post hoc Tukey's adjustment |
| Age of Diagnosis | 0.002 | 0.471 | 0.009 | 9.53E-03 | |
| Male vs Female | -0.154 | 0.163 | -0.127 | 0.252 | |
| LTS(+) & *TERT*(-) vs LTS (-) &*TERT*(-) | -0.592 | 0.201 | -0.625 | 0.175 | 0.363 |
| LTS (-) & *TERT*(+)/Fusion vs LTS(+) & *TERT*(-) | -0.184 | 0.703 | -0.251 | 0.601 | 0.86 |
| LTS (-) & *TERT*(+)/Fusion vs LTS(-) & *TERT*(-) | -0.775 | 6.96E-07 | -0.876 | 9.29E-08 | 2.78E-07 |

**Online-Only References**

1. DeBoy EA, Nicosia AM, Liyanarachchi S, et al. Telomere-lengthening germline variants predispose to a syndromic papillary thyroid cancer subtype. *Am J Hum Genet*. 2024;111(6):1114-1124. doi:10.1016/j.ajhg.2024.04.006

2. Lancaster HO. Significance Tests in Discrete Distributions. *Journal of the American Statistical Association*. 1961;56(294):223-234. doi:10.1080/01621459.1961.10482105

3. Cancer Genome Atlas Research Network. Integrated genomic characterization of papillary thyroid carcinoma. *Cell*. 2014;159(3):676-690. doi:10.1016/j.cell.2014.09.050
